# The gut microbiome is a significant risk factor for future chronic lung disease

**DOI:** 10.1101/2022.03.22.22272736

**Authors:** Yang Liu, Shu Mei Teo, Guillaume Meric, Howard H.F. Tang, Qiyun Zhu, Jon G Sanders, Yoshiki Vazquez-Baeza, Karin Verspoor, Ville A Vartiainen, Pekka Jousilahti, Leo Lahti, Teemu Niiranen, Aki S. Havulinna, Rob Knight, Veikko Salomaa, Michael Inouye

## Abstract

**Background:** The gut-lung axis is generally recognized, but there are few large studies of the gut microbiome and incident respiratory disease in adults.

**Objectives3:** To investigate the associations between gut microbiome and respiratory disease and to construct predictive models from baseline gut microbiome profiles for incident asthma or chronic obstructive pulmonary disease (COPD).

**Methods:** Shallow metagenomic sequencing was performed for stool samples from a prospective, population-based cohort (FINRISK02; N=7,115 adults) with linked national administrative health register derived classifications for incident asthma and COPD up to 15 years after baseline. Generalised linear models and Cox regressions were utilised to assess associations of microbial taxa and diversity with disease occurrence. Predictive models were constructed using machine learning with extreme gradient boosting. Models considered taxa abundances individually and in combination with other risk factors, including sex, age, body mass index and smoking status.

**Results:** A total of 695 and 392 significant microbial associations at different taxonomic levels were found with incident asthma and COPD, respectively. Gradient boosting decision trees of baseline gut microbiome predicted incident asthma and COPD with mean area under the curves of 0.608 and 0.780, respectively. For both incident asthma and COPD, the baseline gut microbiome had C-indices of 0.623 for asthma and 0.817 for COPD, which were more predictive than other conventional risk factors. The integration of gut microbiome and conventional risk factors further improved prediction capacities. Subgroup analyses indicated gut microbiome was significantly associated with incident COPD in both current smokers and non-smokers, as well as in individuals who reported never smoking.

**Conclusions:** The gut microbiome is a significant risk factor for incident asthma and incident COPD and is largely independent of conventional risk factors.

## Introduction

Asthma and chronic obstructive pulmonary disease (COPD) represent the vast majority of chronic respiratory diseases worldwide, causing a considerable burden on health and economy[1, 2]. Both asthma and COPD are recognized as heterogeneous diseases with diverse phenotypes and various underlying mechanisms[3-6]. Currently, spirometry-confirmed airflow limitation is the most common reference standard for establishing diagnoses of asthma and COPD, yet a negative spirometry test result does not rule out the disease[7, 8]. Other criteria that complement evaluation include self-reported symptoms, medical history, physical examination and other diagnoses such as infection, interstitial lung disease, and others[7, 9]. Despite rapidly changing assessments and treatments, both asthma and COPD remain largely underdiagnosed and thus undertreated, leading to lesser quality of life and poorer disease outcomes[3, 9].

With recent advances in high-throughput sequencing, improved characterisation of the human respiratory and gastrointestinal microbiome has been followed by growing recognition of the link between human microbiota and chronic respiratory disease[10, 11]. The gut microbiome is by far the largest and most studied microbial community in the human body[11, 12]. Although the lung microbiome has become well characterized only recently, the link between the lung microbiome and respiratory diseases has been generally acknowledged[10, 13-15]. “Dysbiotic” changes in both airway and gut microbiome have been linked to respiratory diseases; however, the precise mechanism or causal pathway is, as yet, not well understood[16-19]. Emerging evidence suggests cross-talk between gut microbiome and the lungs, via changes to immune responses as well as an interaction of microbiota between the sites, in a hypothesised “gut-lung axis”[11, 20].

Existing studies on the association between gut microbiota and asthma have focused mainly on disease development during childhood[21-23] which is driven by evidence of the influence of early-life microbial exposures on immune function[24, 25]. Previous cross-sectional studies have reported compositional and functional differences of the gut microbiome between adult asthma patients and healthy controls [26-29]. However, little is known about whether and to what extent the gut microbiome affects prospective risk of developing incident asthma in adults. For COPD, there have been far fewer studies on the link between the gut microbiome and disease. Recently, the first analysis of gut microbiome in COPD by Bowerman et al. reported that the faecal microbiome and metabolome significantly differentiate COPD patients and healthy controls[30], which suggests a possible avenue for further investigation using prospective population-scale datasets. Finally, it is only in recent years that methodological and technological advances have opened up the possibility of using large-scale microbial data to predict human respiratory disease[22, 31], but the feasibility of such measures has yet to be evaluated for COPD.

Here we report association analysis and predictive modelling of the gut microbiome and incident asthma and COPD using stool samples from >7,000 participants of a prospective population-based cohort (FINRISK 2002) with electronic health records (EHRs) over ∼15 years of follow-up[32]. Specifically, we (1) describe the gut microbial composition from shallow shotgun metagenomic sequencing and assess the associations with incident asthma and COPD, (2) employ machine learning approaches to quantify the predictive capacities of the gut microbiome at baseline for incident respiratory disease, and (3) construct integrated models of the gut microbiome and conventional risk factors and evaluated their predictive performance.

## Results

A total of 7,115 FINRISK02 participants with baseline gut microbiome profiles and EHR linkage were available for the present study. A summary description of the cohort is given in the **Methods** and baseline characteristics are reported in **Table 1**. After quality control and exclusion criteria were applied, 435 and 145 incident cases of asthma and COPD, respectively, occurred during a median follow-up of 14.8 years after gut microbiome sampling at baseline. Notably, more males than females developed COPD, and incident COPD cases displayed older baseline age than non-cases (P<0.001). The age of onset of incident COPD was significantly older compared to incident asthma (P<0.001). A higher body mass index (BMI) was observed in asthma cases vs non-cases (P=0.002), while there was no significant difference in BMI between COPD cases and non-cases. For both COPD and asthma, a higher proportion of current smokers during the survey year were observed in disease cases than non-cases.

**Table 1.** Characteristics of study participants.

| Characteristic | Asthma |  | COPD |  |
| --- | --- | --- | --- | --- |
|  | Incident cases<br>(n = 435) | Non-cases<br>(n = 5244) | Incident cases<br>(n = 145) | Non-cases<br>(n = 5932) |
| **Females, n (%)** | 252 (57.9%) | 2740 (52.3%) | 43 (29.7%) | 3204 (54%) |
| **Baseline age, years** | 50.9 (40.5-60.5) | 50.5 (39.2-59.3) | 59.5 (53.6-66.5) | 50.5 (39.2-59.5) |
| **Age at first event, years** | 57.6 (46.7-67.3) | -- | 69.1 (61.3-73.6) | -- |
| **Body mass index, kg/m<sup>2</sup>** | 26.7 (24-30.7) | 26.3 (23.7-29.3) | 26.6 (23.5-29.6) | 26.4 (23.7-29.5) |
| **Current smoker, n (%)** | 151 (34.8%) | 1192 (22.8%) | 105 (72.9%) | 1313 (22.2%) |
| **Ex-smoker, n (%)** | 94 (21.6%) | 1181 (22.5%) | 32 (22.1%) | 1321 (22.3%) |

Continuous variables are presented as median (IQR).

### Gut microbiome composition and taxon-level abundances

Individual gut microbiome compositions were characterized by shallow shotgun metagenomic sequencing of stool samples (**Methods**). The present study focused on microbial taxa whose relative abundance exceeded 0.01% in at least 1% of samples; this yielded 46 phyla, 71 classes, 124 orders, 232 families, 617 genera and 1,224 species, as classified according to the Genome Taxonomy Database (GTDB) release 89[33]. The majority of the gut microbiota were dominated by the Firmicutes_A and Bacteroidota phyla (**Fig 1A**), which mostly comprised members of classes Clostridia and Bacteroidia, respectively. At the genus level, *Faecalibacterium* and *Agathobacter* in phylum Firmicutes_A, as well as *Bacteroides, Bacteroides_B* and *Prevotella* in phylum Bacteroidota were most abundant in a majority of samples (**Fig 1B**).

**Fig 1.**
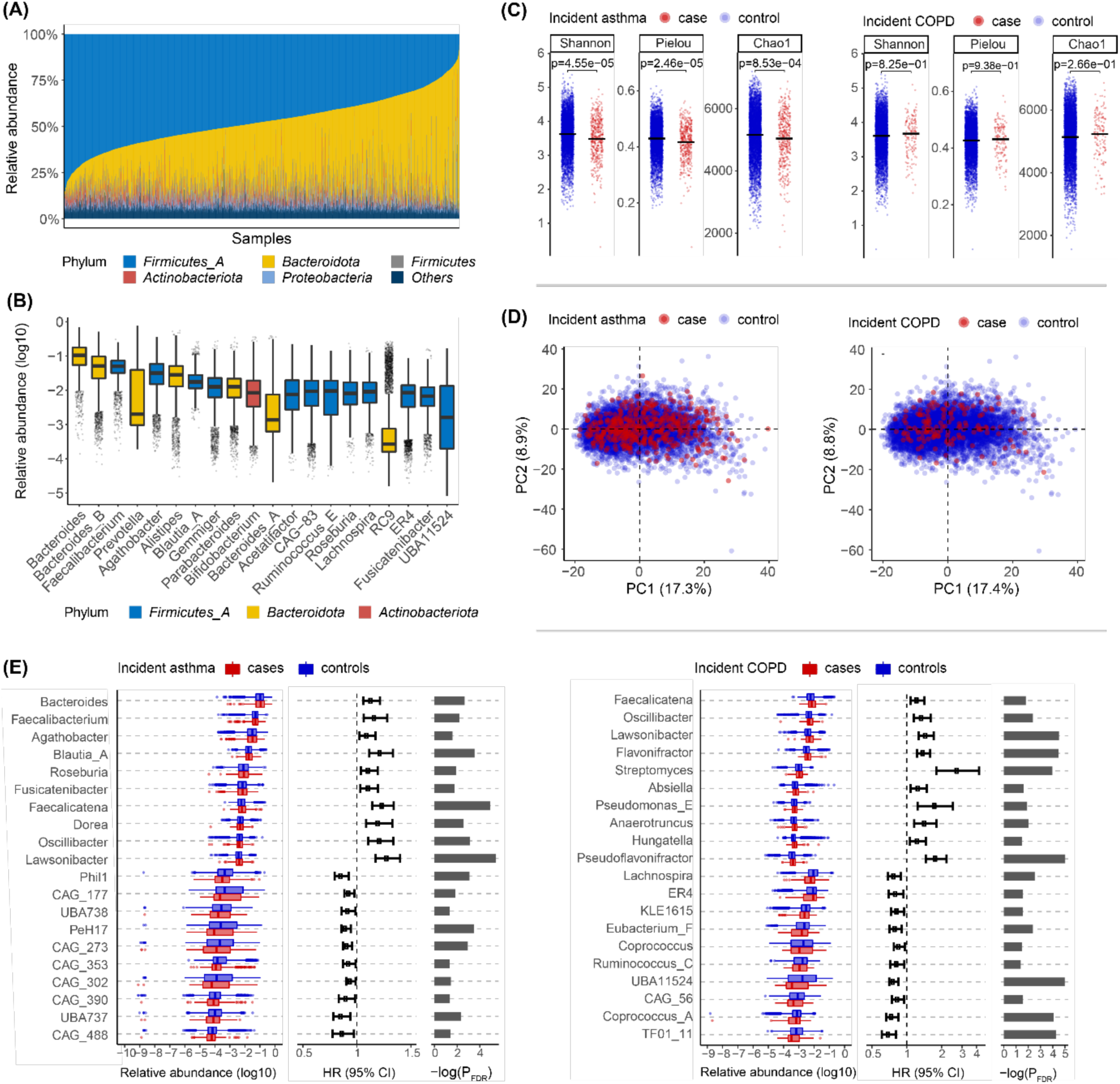
Gut microbiome composition and characteristics. **A**, Gut microbiome profiles at phylum level. **B**, Box plots of the 20 most abundant genera sorted by mean relative abundance. **C**, Shannon’s, Pielous’s and Chao1 indices at genus level between cases and non-cases. Median values are represented by horizontal lines. **D**, Principal component analysis on centered log-ratio transformed abundances at genus level. **E**, Genera associated with incident asthma or COPD surpassing a false discovery rate threshold of 5% (P_FDR_<0.05). Only the top 10 most abundant genera for each of combination of positive or negative associations, with COPD or asthma.

Baseline alpha-diversity measures significantly differed between incident asthma cases and non-cases (P<0.01), with lower values of Shannon’s, Chao1, and Pielou’s indices in individuals who went on to develop asthma (**Fig 1C**). Alpha-diversity indices were not significantly different between COPD cases and non-cases. Principal component analysis of the centered log-ratio (CLR) transformed abundances showed no clear separation between incident cases and non-cases (**Fig 1D**), suggesting that the association of incident asthma and COPD with the gut microbiome was unlikely related to the whole microbial community and may be attributable to specific microbial taxa.

We assessed the association between baseline taxon-level microbial abundances and incident respiratory diseases using Cox regression, based on centered log-ratios (**Methods**). At 5% false discovery rate, significant associations of incident asthma were found in 5 phyla, 5 classes, 18 orders, 111 families, 257 genera and 299 species (**Table S1**); for incident COPD, we found significant associations with 5 phyla, 7 classes, 32 orders, 57 families, 133 genera and 158 species (**Table S2**). Of the asthma-and COPD-associated taxa, 76% and 68.6% showed positive associations with disease incidence, respectively. A number of highly abundant genera were associated with incident asthma, such as *Bacteroides, Faecalibacterium, Agathobacter, Blautia_A* and *Roseburia* (**Fig 1E**). Among the most abundant COPD-associated genera, increased abundance of *Faecalicatena, Oscillibacter, Lawsonibacter, Flavonifractor* and *Streptomyces*, and reduced abundances of *Lachnospira, ER4, KLE1615, Eubacterium_F* and *Coprococcus* were associated with incident COPD.

### Gut microbiome and gradient boosting decision trees to predict incident asthma and COPD

To investigate whether the baseline gut microbiome was predictive of incident asthma and COPD, we train and validate prediction models via the machine learning algorithm of gradient boosting decision trees. These models were trained with 5-fold cross-validation in 70% of the individuals and then the performances were validated in the remaining 30% (**Methods**); all performance metrics given are based on the 30% validation set unless otherwise specified. Models were developed at different taxonomic levels separately and for a combination of all taxonomic levels (**Fig S1**). To assess sampling variation, we resampled training and testing partitions at different taxonomic levels 10 times and report mean values of prediction performance.

The best performance was obtained at individual taxonomic levels, rather than their combination, for both asthma and COPD prediction. Generally better prediction performance was attained at lower taxonomic levels, particularly for COPD where the highest average area under the operating characteristic curve (AUC) was at species level (mean AUC = 0.780), followed by genus (mean AUC = 0.734) and family (mean AUC = 0.688) levels. For prediction of incident asthma, the best performance was obtained at family level (mean AUC = 0.608), with slight attenuation of AUC scores obtained at genus (mean AUC = 0.592) and species (mean AUC = 0.593) levels.

### The gut microbiome had greater predictive value than individual conventional risk factors

To compare the predictive value of conventional risk factors and the gut microbiome for incident asthma and COPD, we first conducted univariate analysis using Cox models. We utilised the optimal cross-validated gradient boosting model at family and species level for asthma and COPD, respectively, and refer to the resultant score as a “gut microbiome score” for each condition. We found that the gut microbiome score had a relatively high predictive capacity with C-indices of 0.623 for asthma and 0.817 for COPD, which were each greater than those of other risk factors (**Fig 2**). Smoking status at baseline was significantly associated with increased risk of both asthma (HR=2.21, 95% CI [1.53-3.20], P <0.001) and COPD (HR=8.16 [4.55-14.64], P<0.001) compared with non-smoking (**Table 2**). Increased incidence of COPD was also significantly associated with male sex (HR=2.19 [1.25-3.82], P=0.01) and older baseline age (HR=1.07 per year, [1.04-1.10], P<0.001). The gut microbiome score was associated with increased incidence of both asthma (HR=1.44 per s.d., [1.23-1.67], P<0.001) and COPD (HR=1.39 per s.d., [1.30-1.49], P<0.001).

**Table 2.**
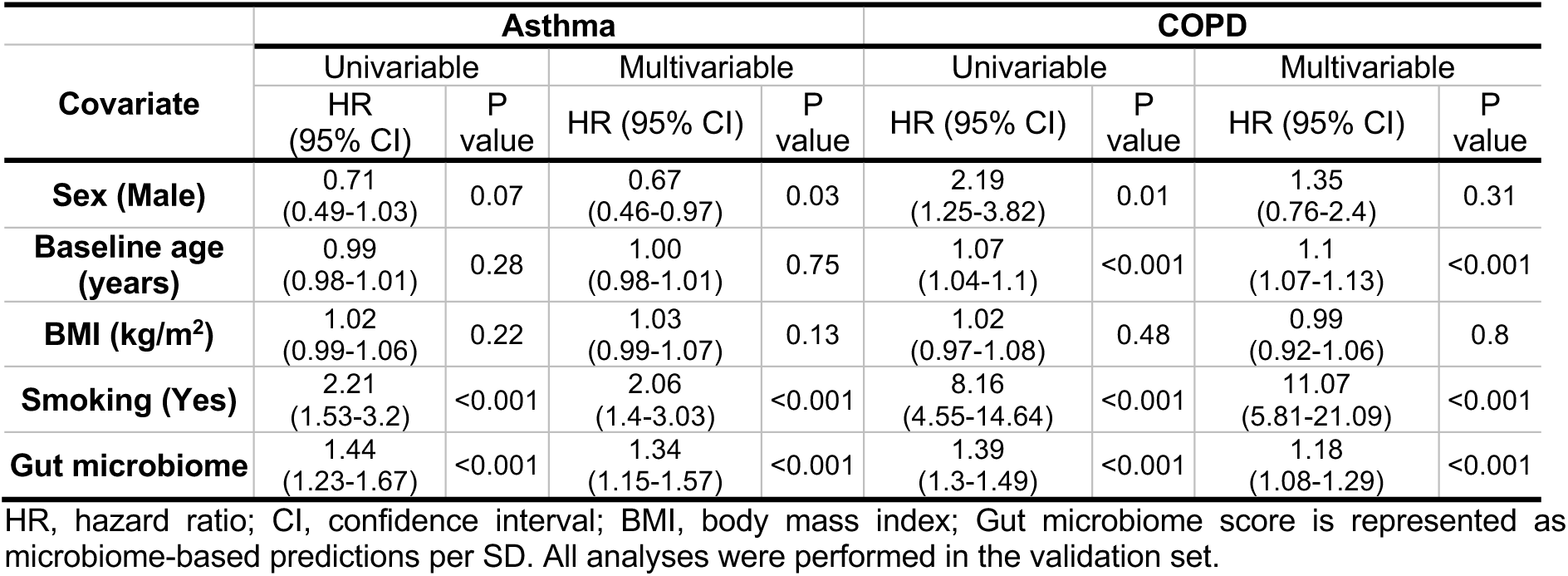
Association of risk factors separately and combined for incident asthma and COPD.

| Covariate | Asthma |  |  |  | COPD |  |  |  |
| --- | --- | --- | --- | --- | --- | --- | --- | --- |
|  | Univariable |  | Multivariable |  | Univariable |  | Multivariable |  |
|  | HR<br>(95% CI) | P<br>value | HR (95% CI) | P<br>value | HR (95% CI) | P<br>value | HR (95% CI) | P<br>value |
| **Sex (Male)** | 0.71<br>(0.49-1.03) | 0.07 | 0.67<br>(0.46-0.97) | 0.03 | 2.19<br>(1.25-3.82) | 0.01 | 1.35<br>(0.76-2.4) | 0.31 |
| **Baseline age**<br>**(years)** | 0.99<br>(0.98-1.01) | 0.28 | 1.00<br>(0.98-1.01) | 0.75 | 1.07<br>(1.04-1.1) | <0.001 | 1.1<br>(1.07-1.13) | <0.001 |
| **BMI (kg/m<sup>2</sup>)** | 1.02<br>(0.99-1.06) | 0.22 | 1.03<br>(0.99-1.07) | 0.13 | 1.02<br>(0.97-1.08) | 0.48 | 0.99<br>(0.92-1.06) | 0.8 |
| **Smoking (Yes)** | 2.21<br>(1.53-3.2) | <0.001 | 2.06<br>(1.4-3.03) | <0.001 | 8.16<br>(4.55-14.64) | <0.001 | 11.07<br>(5.81-21.09) | <0.001 |
| **Gut microbiome** | 1.44<br>(1.23-1.67) | <0.001 | 1.34<br>(1.15-1.57) | <0.001 | 1.39<br>(1.3-1.49) | <0.001 | 1.18<br>(1.08-1.29) | <0.001 |

HR, hazard ratio; CI, confidence interval; BMI, body mass index; Gut microbiome score is represented as microbiome-based predictions per SD. All analyses were performed in the validation set.

**Fig 2.**
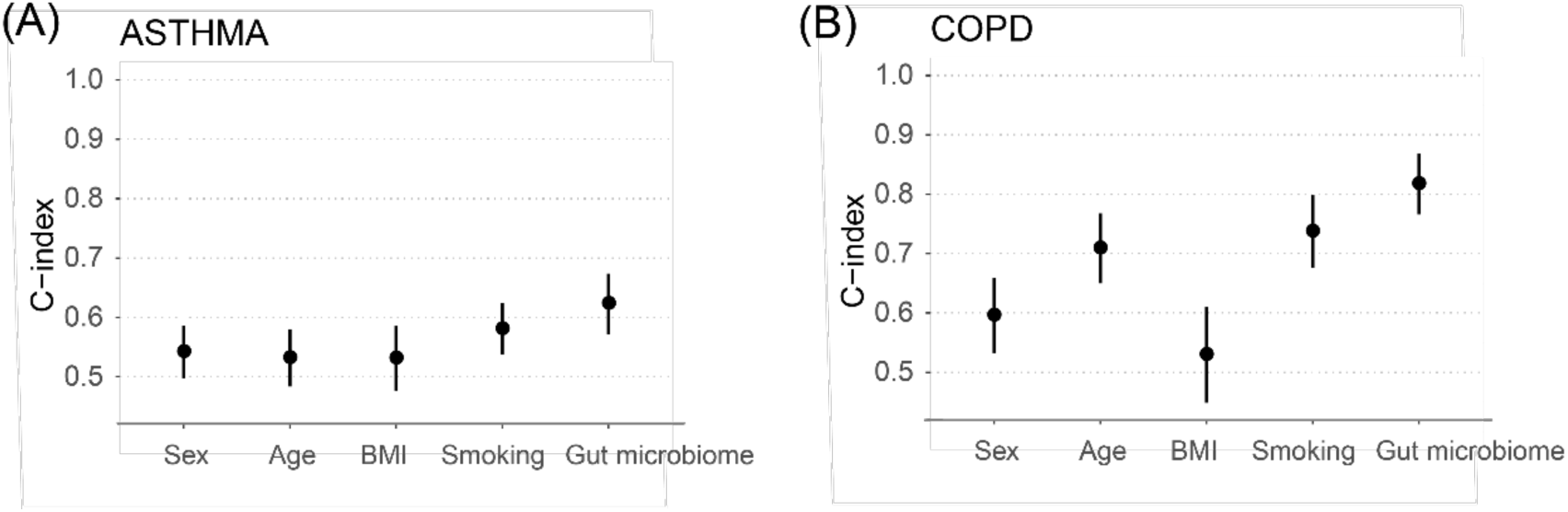
Predictive capacity of each risk factor separately for **A**, incident asthma or **B**, COPD. Univariate Cox models were performed for each of sex, baseline age, BMI, smoking and gut microbiome individually. Points and error bars represent the C-indices and 95% confidence intervals.

### Integrated prediction models of the gut microbiome and conventional risk factors

When integrating risk factors and gut microbiome score, the Cox model for asthma showed that current smoking status and gut microbiome were significantly associated with higher risk (HR=2.06 [1.40-3.03], P<0.001, and HR=1.34 per SD [1.15-1.57], P<0.001, respectively), and male sex was significantly associated with lower risk (HR=0.67 [0.46-0.97], P=0.03), whereas there were no significant associations for baseline age and BMI (**Table 2**). For COPD, baseline age, current smoking status and gut microbiome score were significant predictors (HR= 1.1 per year [1.07-1.13] P<0.001; HR=11.07 [5.81-21.09], P<0.001; and HR=1.18 per SD [1.08-1.29], P<0.001 respectively). While consistent with the individual predictive power of the gut microbiome score (**Fig 2**), the multivariable Cox model showed the risk associated with current smokers at baseline was significantly greater than other risk factors for COPD.

In subgroup analyses, the gut microbiome score association patterns were generally consistent with those above (**Fig 3**). For COPD, where current smoking status had a relatively large hazard ratio, the gut microbiome score was independently associated with incident COPD in both current smokers and non-smokers. In individuals who indicated past smoking but who were not current smokers at survey (n=414), we found that the gut microbiome score was not significantly associated with incident COPD (HR=1.22 [0.89-1.68], P=0.22) but that, in individuals who reported never smoking (n=970), there was a significant association with incident COPD (HR=1.40 [1.02-1.91], P=0.04). Finally, in COPD, we observed evidence for statistical interactions of the gut microbiome score with age and sex (**Fig 3**).

**Fig 3.**
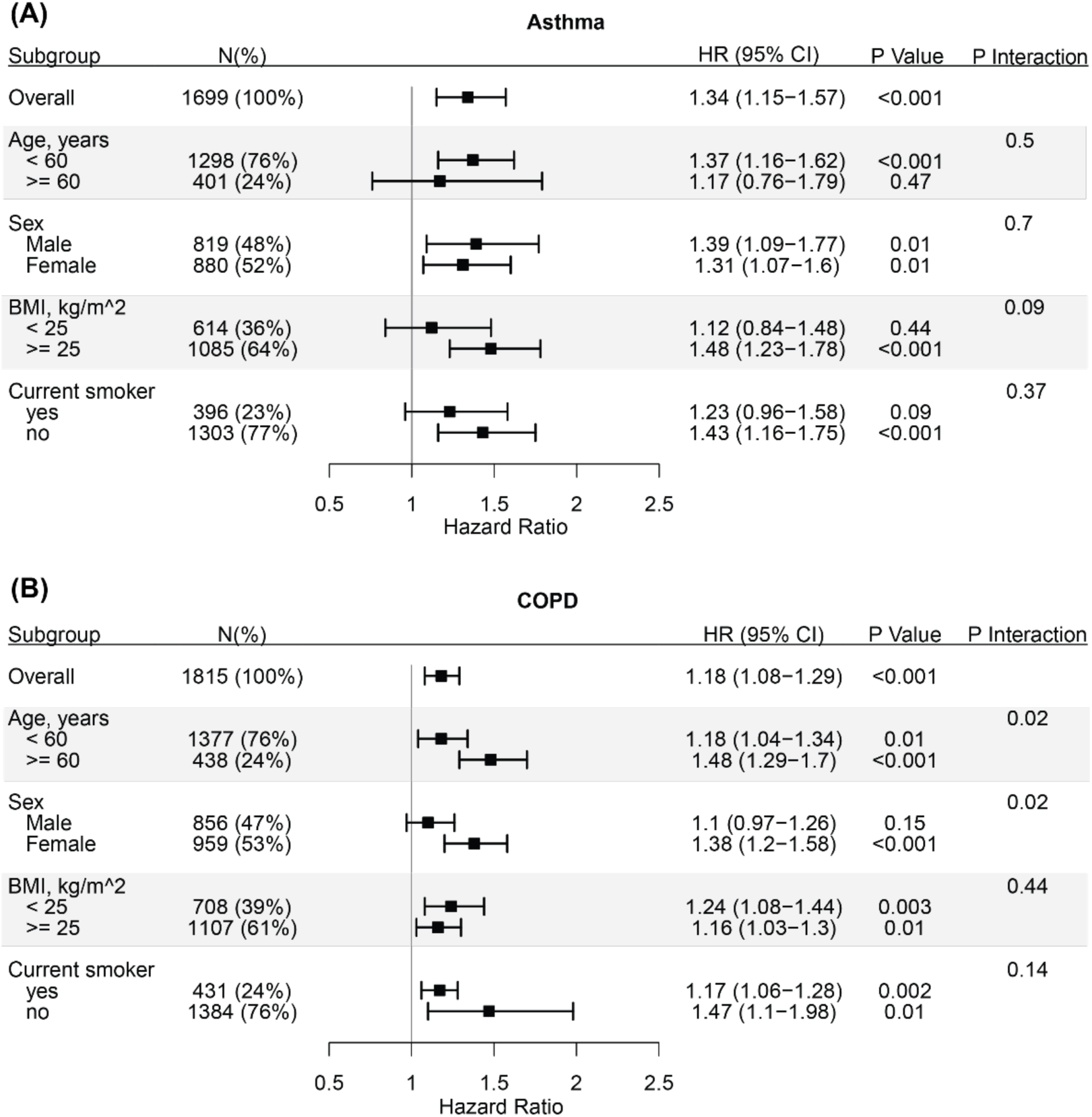
Subgroup analyses for **A**, incident asthma or **B**, COPD. Cox models were applied to test for interactions between gut microbiome and patient characteristic subgroups. Points and error bars represent hazard ratios per SD and 95% confidence intervals of gut microbiome score across subgroups.

The integrated models showed significantly improved predictive capacity for both incident asthma and COPD (**Fig 4**). For asthma, a reference model of age, sex and BMI yielded C-index of 0.567; addition of smoking status then gut microbiome score increased the C-index further to 0.626 and 0.656, respectively. For COPD, the reference model of age, sex and BMI yielded C-index of 0.735; addition of smoking status then gut microbiome score increased the C-index further to 0.855 and 0.862, respectively.

**Fig 4.**
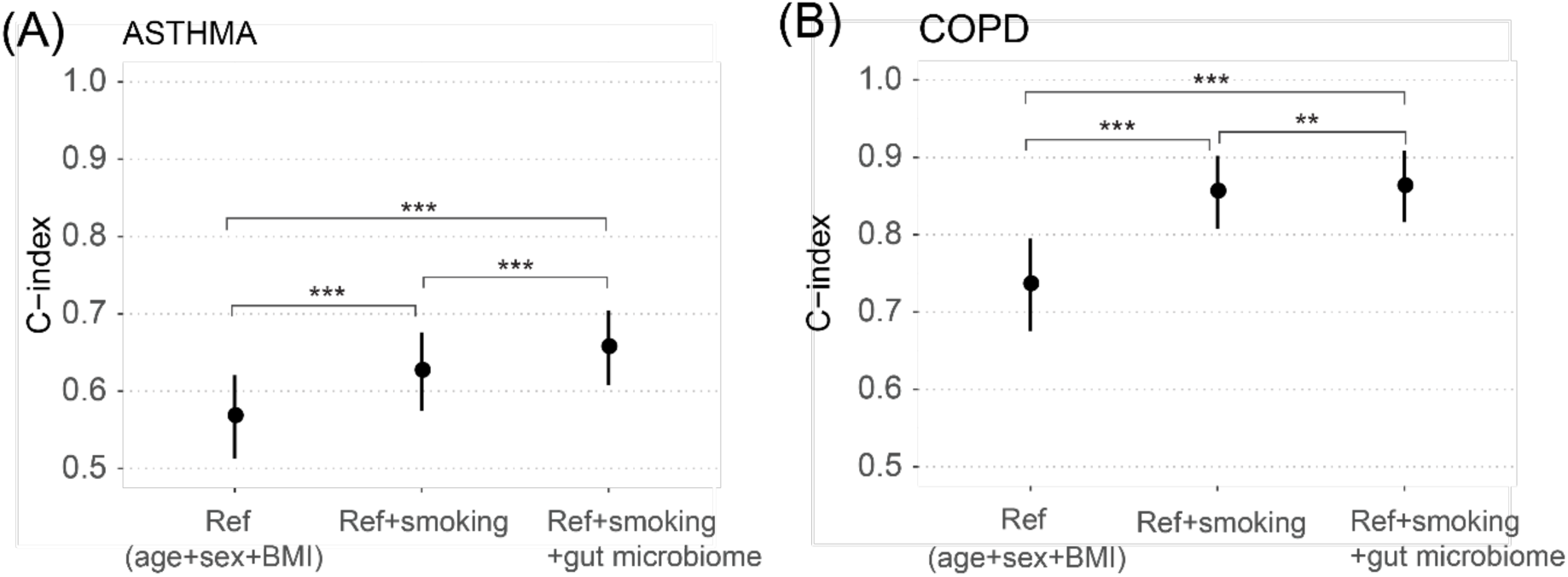
Predictive capacity of integrated models for **A**, incident asthma and **B**, COPD. ‘Ref’ is a reference model that jointly considers age, sex and BMI. Points and error bars represent the C-indices and 95% confidence intervals. Analysis of deviance based on the log partial likelihood, P<0.01, **; P<0.001, ***.

## Discussion

In this prospective study, we investigated the association and predictive capacity of the gut microbiome for future chronic respiratory diseases, asthma and COPD, in adults using shotgun metagenomics. We demonstrated that the gut microbiome is significantly associated with incident asthma and COPD and evaluated the relative contributions of traditional risk factors and a gut microbiome score. We then constructed integrated risk models which maximised predictive performance. Taken together, our findings indicate that the gut microbiome is a valid and potentially substantive biomarker for both asthma and COPD.

The gut and lung microbial communities, although residing in distal sites, are dominated by broadly similar bacterial phyla, including Firmicutes and Bacteroidetes, but differ in local compositions and total microbial biomass[11]. Some of our findings are relevant to previous microbial studies of the respiratory tract. For example, *Haemophilus* and *Streptococcus* have been previously found to be positively associated with respiratory illnesses in the airways [18, 34, 35]. In our gut microbiome samples, we also found positive associations between *Streptococcus* and incident asthma; however, we found that multiple *Haemophilus spp*. were significantly negatively associated with incident COPD. An increased abundance of *Pseudomonas spp*. from the airway microbiome was previously reported in COPD exacerbations[36, 37] and impaired pulmonary function[38, 39]. Consistent with this, we found positive associations of the *Pseudomonas, Pseudomonas_A* and *Pseudomonas_E* genera (all part of *Pseudomonas* according to the NCBI taxonomy) with incident asthma and COPD. These findings support the emerging evidence of possible functional links between the respiratory tract and gastrointestinal tract, however the underlying mechanisms by which microorganisms between the sites may interact remain unclear[40, 41].

Despite increasing recognition of the existence of gut-lung crosstalk, the role of the gut microbiota in respiratory disease has been primarily studied in children. Its relevance in adults has been unclear. Previous studies have demonstrated that the early-life gut microbial alteration and maturation patterns influence the risk of asthma development in childhood[22, 23, 42]. In our data, we found that higher abundances of *Escherichia[31], Enterococcus, Clostridium, Veillonella*, and *B. fragilis* were associated with increased incidence of asthma in adulthood, consistent with that observed for childhood asthma[22, 43, 44]. In contrast to previous findings showing that the relative abundances of *Faecalibacterium, Roseburia* and *Flavonifractor* were decreased in childhood asthma[22, 43], we found positive associations with adult-onset asthma. We confirmed previous findings that increased abundances of *Clostridium* and *Eggerthella lenta* in the adult gut microbiome were associated with asthma[27]. The relationship between the gut microbiome and COPD is even less understood. A recent study reported that *Streptococcus sp000187445* was enriched in COPD patients and was correlated with reduced lung function[30], which was also confirmed by a positive association with incident COPD in our study.

Regarding consideration of causality in observational studies, it is challenging to determine whether the composition of the gut microbiome is a cause or consequence of respiratory disease. In this respect, one strength of our study was the use of baseline gut microbiome and incident disease systematically identified through EHRs. The follow-up using EHRs was nearly complete in all samples (except for the small number of participants who moved abroad permanently). Using machine learning models, we found that the baseline gut microbiome had moderate predictive capacities in distinguishing incident cases from non-cases for asthma and COPD, suggesting that there are detectable changes in the gut microbiome antecedent to the onset of symptomatic disease. This does not confirm causality or eliminate other possibilities. For example, disease-associated host changes and gut microbial alteration may influence each other and operate simultaneously[40]. We also showed that the association between gut microbiome-based predictions and incident asthma or COPD was largely independent of age, sex, BMI and smoking, all of which can influence susceptibility to respiratory diseases[45-48]. Moreover, significant interactions of gut microbiome by sex and age were found, suggesting different impact of gut microbiome on age and sex groups, consistent with findings in other settings[49-51].

Importantly, our study affirms the large body of evidence that smoking is associated with respiratory illness, especially COPD. Despite many ways to characterise the smoking phenotype, we found that individuals who reported being current smokers were at high risk of future asthma and COPD. The association between smoking and gut microbiota is well established and smoking cessation has been shown to have profound, putatively causal effects on the gut microbiome[52]. Our results show that, particularly for COPD, the gut microbiome is both a substantial independent predictor of future disease and that its predictive power is partially explained by smoking behaviour. As such, our findings are both consistent with previous studies and take us a step closer to delineating which and to what extent particular gut microbial taxa sit along the causal path from smoking behaviour to future asthma and COPD. For the latter, larger prospective studies will be necessary but population-scale gut microbiome and e-health studies are under way.

There are limitations of the present study. Firstly, despite a relatively large sample size, our study was enrolled from a single European country (Finland), and the generalizability of the findings to other geographically- and culturally-distinct settings will require further investigation. Furthermore, only one time point of the gut microbiome was sampled per individual, which did not allow for dynamic or temporal assessment of gut microbiome alterations along with incident disease onset. Changes in diet and environmental exposures (apart from smoking) can induce changes in gut microbiota and should be considered in future studies. While the asthma and COPD phenotypes can be difficult to diagnosis or indeed overlap in some individuals, our study takes a pragmatic approach and future clinical cohorts may be necessary to precisely quantify disease specific effects. Finally, although formal lung function test results (FEV1, FVC) may further improve prediction, it was not feasible to perform wholescale clinical examination of airflow obstruction at the population level. Regardless, our study demonstrates that future exploration of the influence of the gut microbiome in severity and progression of asthma and COPD is warranted, and may lead to further clinically-significant findings.

Our study supports the role of gut microbiome in adult respiratory disease and as potential biomarkers that might aid in risk profiling of asthma and COPD. The underlying mechanisms and causal links by which gut microbiota influence the lung, and vice versa, remain to be established.

## Methods

### Study design and participants

The FINRISK 2002 study is a population-based nationwide survey carried out in Finland in 2002, consisting of random samples of the population aged 25 to 74 years drawn from the National Population Information System[32]. The survey included self-administered questionnaires, health examinations conducted at the study sites by trained personnel, and collection of biological samples. The overall participation rate was 65.5% (n = 8798). The participants were followed up through linkage to national administrative electronic registers that proved highly reliable[53-55]. Inclusion criteria have been described elsewhere[32]. Exclusion criteria for the present study are missing follow-ups, prior diagnosis of the disease for prediction, baseline pregnancy, systemic use of antibiotics at baseline and unmet sequencing depth. The incident cases of asthma and COPD were identified according to ICD-10 diagnosis codes (Finnish modification) from linked EHRs which were last followed up by Dec 31^st^, 2016. COPD cases were defined using ICD codes J43|J44; asthma cases were defined using ICD codes J45|J46, or the Social Insurance Institution of Finland (Kela) reimbursement code 203 for asthma medication, or medicine purchases with ATC codes R03BA|R03BC|R03DC|R03AK. Covariates included baseline age, sex, body mass index (BMI), and smoking. Written informed consent was obtained from all participants. The Coordinating Ethics Committee of the Helsinki and Uusimaa Hospital District approved the FINRISK 2002 study protocols (Ref. 558/E3/2001). The study was conducted according to the World Medical Association’s Declaration of Helsinki on ethical principles.

### Sample collection

During the baseline survey, stool samples were collected by participants at home using a collection kit with instructions, and mailed overnight under winter conditions to the Finnish Institute for Health and Welfare for storing at -20°C. The frozen stool samples were transferred to University of California San Diego for sequencing in 2017.

### DNA extraction, sequence processing and taxonomic profiling

The gut microbiome was characterized by shallow shotgun metagenomics sequencing[56] with an Illumina HiSeq 4000 platform to a mean depth of ∼10^6^ reads/sample. The stool shotgun sequencing was successfully performed in 7,231 individuals. Libraries were prepared using KAPA HyperPlus Kit according to manufacturer’s protocol. Sequencing reads were processed using the Snakemake pipeline[57]. Removal of low quality, adapter and host reads was performed. The details of DNA extraction and library preparation for stool samples have been described elsewhere[31]. Samples were filtered by sequencing depth of 400,000 reads/sample to preserve data quality and the majority of disease cases which resulted in 7163 samples remaining. The metagenomes were classified using default parameters in Centrifuge 1.0.4[58], and using an index database based on taxonomic definitions from the Genome Taxonomy Database (GTDB) release 89[33]. In total, 151 phyla, 338 classes, 925 orders, 2,254 families, 7,906 genera and 24,705 species were uniquely identified based on GTDB taxonomy. The relative abundances of bacterial taxa at phylum, class, order, family, genus and species levels were computed. The present analyses focused on common taxa with relative abundances greater than 0.01% in more than 1% of samples. Three measures of microbial diversity were calculated: Shannon’s alpha diversity, Chao1 richness and Pielou’s evenness (R packages vegan and otuSummary). The centered log-ratio (CLR) transformation was performed on abundance data, of which the zeros were substituted with 1/10 of non-zero minimum abundance. Further analyses were based on CLR transformed abundances.

### Machine learning and statistical analysis

A machine learning framework was employed to develop prediction models at different taxonomic levels separately. The samples were randomly partitioned into two subsets: (1) a training dataset (70% of samples) for developing models, and (2) a validation dataset (30% of samples) for evaluating prediction performance. We resampled the data 10 times and performed the same training and validation procedure for each sampling partition. In each training dataset, we first selected microbial indicators for predicting incident asthma and COPD; we analyzed the relationships between taxon-level abundance and incident disease using logistic regression adjusted for age and sex, Cox regression for time to disease onset adjusted for age and sex and Spearman correlation. The taxa that were associated with incident diseases at a significance threshold of P<0.05 by any of the above approaches were selected for further analyses. The selected taxa together with diversity measurements were considered as microbial predictors for developing prediction models. Next, gradient boosting decision tree (implemented by Xgboost) models were developed with Bayesian optimization through 5-fold cross-validation to determine optimal hyperparameters. The optimal setting was then trained on the whole training data to build the final model used in validation. We additionally performed ridge logistic regression to compare the prediction performance using the same samples for training and testing. The gradient boosted trees-based models outperformed those based on ridge logistic regression. A similar trend of prediction performance across taxonomic levels was observed with both methods. The final performance across various models and partitions was assessed in the validation datasets.

Wilcoxon rank-sum test was performed to compare differences in patient characteristics, gut microbial relative abundances and diversity metrics between incident cases and non-cases. Cox regression with adjustment of age and sex was utilized to assess the association between taxon-level CLR abundance and incident disease using all samples (FDR<0.05 was considered as statistical significance). The gut microbiome-based predictions from the optimal gradient boosting model were used as the gut microbiome scores for further analyses in its respective validation dataset for each disease condition. Cox models of conventional risk factors and in combination with the gut microbiome score were built using the time from baseline to the occurrence of the disease or end of follow-up. Machine learning and statistical analysis of data were carried out in R (version 3.6.1).

## Data Availability

The FINRISK data for this study are available with a written application to the THL Biobank as instructed on the website: https://thl.fi/en/web/thl-biobank/for-researchers.

## Data and code availability

The FINRISK data for this study are available with a written application to the THL Biobank as instructed on the website: https://thl.fi/en/web/thl-biobank/for-researchers. A separate permission is needed from FINDATA (www.findata.fi/en/) for use of the EHR data. Custom code for analysis in this study is available at https://github.com/yangl700/microb_pred.

## Acknowledgements

VS was supported by the Finnish Foundation for Cardiovascular Research and by Juho Vainio Foundation. MI was supported by the Munz Chair of Cardiovascular Prediction and Prevention. ASH was supported by the Academy of Finland, grant no. 321356. LL was supported by Academy of Finland (295741, 328791). TN was supported by the Emil Aaltonen Foundation, the Finnish Foundation for Cardiovascular Research and the Academy of Finland (grant no. 321351). This study was supported by the Victorian Government’s Operational Infrastructure Support (OIS) program and by core funding from the British Heart Foundation (RG/13/13/30194; RG/18/13/33946) and the NIHR Cambridge Biomedical Research Centre (BRC-1215-20014) [*]. *The views expressed are those of the author(s) and not necessarily those of the NIHR or the Department of Health and Social Care. This work was supported by Health Data Research UK, which is funded by the UK Medical Research Council, Engineering and Physical Sciences Research Council, Economic and Social Research Council, Department of Health and Social Care (England), Chief Scientist Office of the Scottish Government Health and Social Care Directorates, Health and Social Care Research and Development Division (Welsh Government), Public Health Agency (Northern Ireland), British Heart Foundation and Wellcome.

*The views expressed are those of the authors and not necessarily those of the NHS or the Department of Health and Social Care.

## References

1. Halpin, D.M.G., et al., Global Initiative for the Diagnosis, Management, and Prevention of Chronic Obstructive Lung Disease. The 2020 GOLD Science Committee Report on COVID-19 and Chronic Obstructive Pulmonary Disease. Am J Respir Crit Care Med, 2021. 203(1): p. 24–36.

2. Asthma, G.I.f., Global Strategy for Asthma Management and Prevention. 2020.

3. Papi, A., et al., Asthma. Lancet, 2018. 391(10122): p. 783–800.

4. Barnes, P.J., Therapeutic approaches to asthma-chronic obstructive pulmonary disease overlap syndromes. J Allergy Clin Immunol, 2015. 136(3): p. 531–45.

5. Mirza, S. and R. Benzo, Chronic Obstructive Pulmonary Disease Phenotypes: Implications for Care. Mayo Clin Proc, 2017. 92(7): p. 1104–1112.

6. Kuruvilla, M.E., F.E. Lee, and G.B. Lee, Understanding Asthma Phenotypes, Endotypes, and Mechanisms of Disease. Clin Rev Allergy Immunol, 2019. 56(2): p. 219–233.

7. McCracken, J.L., et al., Diagnosis and Management of Asthma in Adults: A Review. JAMA, 2017. 318(3): p. 279–290.

8. Polverino, F. and B. Celli, The Challenge of Controlling the COPD Epidemic: Unmet Needs. Am J Med, 2018. 131(9S): p. 1–6.

9. Riley, C.M. and F.C. Sciurba, Diagnosis and Outpatient Management of Chronic Obstructive Pulmonary Disease: A Review. JAMA, 2019. 321(8): p. 786–797.

10. Budden, K.F., et al., Functional effects of the microbiota in chronic respiratory disease. Lancet Respir Med, 2019. 7(10): p. 907–920.

11. Chotirmall, S.H., et al., Microbiomes in respiratory health and disease: An Asia-Pacific perspective. Respirology, 2017. 22(2): p. 240–250.

12. Shreiner, A.B., J.Y. Kao, and V.B. Young, The gut microbiome in health and in disease. Curr Opin Gastroenterol, 2015. 31(1): p. 69–75.

13. Huffnagle, G.B., R.P. Dickson, and N.W. Lukacs, The respiratory tract microbiome and lung inflammation: a two-way street. Mucosal Immunol, 2017. 10(2): p. 299–306.

14. Yatera, K., S. Noguchi, and H. Mukae, The microbiome in the lower respiratory tract. Respir Investig, 2018. 56(6): p. 432–439.

15. Wang, Z., et al., Lung microbiome dynamics in COPD exacerbations. Eur Respir J, 2016. 47(4): p. 1082–92.

16. Huang, Y.J., et al., The airway microbiome in patients with severe asthma: Associations with disease features and severity. J Allergy Clin Immunol, 2015. 136(4): p. 874–84.

17. Huang, Y.J., et al., Airway microbiome dynamics in exacerbations of chronic obstructive pulmonary disease. J Clin Microbiol, 2014. 52(8): p. 2813–23.

18. Teo, S.M., et al., The infant nasopharyngeal microbiome impacts severity of lower respiratory infection and risk of asthma development. Cell Host Microbe, 2015. 17(5): p. 704–15.

19. Teo, S.M., et al., Airway Microbiota Dynamics Uncover a Critical Window for Interplay of Pathogenic Bacteria and Allergy in Childhood Respiratory Disease. Cell Host Microbe, 2018. 24(3): p. 341–352 e5.

20. Dang, A.T. and B.J. Marsland, Microbes, metabolites, and the gut-lung axis. Mucosal Immunol, 2019. 12(4): p. 843–850.

21. Barcik, W., et al., The Role of Lung and Gut Microbiota in the Pathology of Asthma. Immunity, 2020. 52(2): p. 241–255.

22. Stokholm, J., et al., Maturation of the gut microbiome and risk of asthma in childhood. Nat Commun, 2018. 9(1): p. 141.

23. Depner, M., et al., Maturation of the gut microbiome during the first year of life contributes to the protective farm effect on childhood asthma. Nat Med, 2020. 26(11): p. 1766–1775.

24. Huang, Y.J. and H.A. Boushey, The microbiome in asthma. J Allergy Clin Immunol, 2015. 135(1): p. 25–30.

25. Tamburini, S., et al., The microbiome in early life: implications for health outcomes. Nat Med, 2016. 22(7): p. 713–22.

26. Barcik, W., et al., Histamine-secreting microbes are increased in the gut of adult asthma patients. J Allergy Clin Immunol, 2016. 138(5): p. 1491–1494 e7.

27. Wang, Q., et al., A metagenome-wide association study of gut microbiota in asthma in UK adults. BMC Microbiol, 2018. 18(1): p. 114.

28. Begley, L., et al., Gut microbiota relationships to lung function and adult asthma phenotype: a pilot study. BMJ Open Respir Res, 2018. 5(1): p. e000324.

29. Hevia, A., et al., Allergic Patients with Long-Term Asthma Display Low Levels of Bifidobacterium adolescentis. PLoS One, 2016. 11(2): p. e0147809.

30. Bowerman, K.L., et al., Disease-associated gut microbiome and metabolome changes in patients with chronic obstructive pulmonary disease. Nat Commun, 2020. 11(1): p. 5886.

31. Salosensaari, A., et al., Taxonomic signatures of cause-specific mortality risk in human gut microbiome. Nat Commun, 2021. 12(1): p. 2671.

32. Borodulin, K., et al., Cohort Profile: The National FINRISK Study. Int J Epidemiol, 2018. 47(3): p. 696–696i.

33. Parks, D.H., et al., A standardized bacterial taxonomy based on genome phylogeny substantially revises the tree of life. Nat Biotechnol, 2018. 36(10): p. 996–1004.

34. Hufnagl, K., et al., Dysbiosis of the gut and lung microbiome has a role in asthma. Semin Immunopathol, 2020. 42(1): p. 75–93.

35. O’Dwyer, D.N., R.P. Dickson, and B.B. Moore, The Lung Microbiome, Immunity, and the Pathogenesis of Chronic Lung Disease. J Immunol, 2016. 196(12): p. 4839–47.

36. Millares, L., et al., Bronchial microbiome of severe COPD patients colonised by Pseudomonas aeruginosa. Eur J Clin Microbiol Infect Dis, 2014. 33(7): p. 1101–11.

37. Garcia-Vidal, C., et al., Pseudomonas aeruginosa in patients hospitalised for COPD exacerbation: a prospective study. Eur Respir J, 2009. 34(5): p. 1072–8.

38. Garcia-Clemente, M., et al., Impact of Pseudomonas aeruginosa Infection on Patients with Chronic Inflammatory Airway Diseases. J Clin Med, 2020. 9(12).

39. Davies, G., et al., The effect of Pseudomonas aeruginosa on pulmonary function in patients with bronchiectasis. Eur Respir J, 2006. 28(5): p. 974–9.

40. Budden, K.F., et al., Emerging pathogenic links between microbiota and the gut-lung axis. Nat Rev Microbiol, 2017. 15(1): p. 55–63.

41. Zhang, D., et al., The Cross-Talk Between Gut Microbiota and Lungs in Common Lung Diseases. Front Microbiol, 2020. 11: pp. 301.

42. Arrieta, M.C., et al., Early infancy microbial and metabolic alterations affect risk of childhood asthma. Sci Transl Med, 2015. 7(307): p. 307ra152.

43. Chiu, C.Y., et al., Gut microbial-derived butyrate is inversely associated with IgE responses to allergens in childhood asthma. Pediatr Allergy Immunol, 2019. 30(7): p. 689–697.

44. Vael, C., et al., Early intestinal Bacteroides fragilis colonisation and development of asthma. BMC Pulm Med, 2008. 8: pp. 19.

45. Han, M.K., et al., Gender and chronic obstructive pulmonary disease: why it matters. Am J Respir Crit Care Med, 2007. 176(12): p. 1179–84.

46. Zein, J.G. and S.C. Erzurum, Asthma is Different in Women. Curr Allergy Asthma Rep, 2015. 15(6): p. 28.

47. Zammit, C., et al., Obesity and respiratory diseases. Int J Gen Med, 2010. 3: p. 335–43.

48. Sears, M.R., Smoking, asthma, chronic airflow obstruction and COPD. Eur Respir J, 2015. 45(3): p. 586–8.

49. O’Toole, P.W. and I.B. Jeffery, Gut microbiota and aging. Science, 2015. 350(6265): p. 1214–5.

50. Haro, C., et al., Intestinal Microbiota Is Influenced by Gender and Body Mass Index. PLoS One, 2016. 11(5): p. e0154090.

51. Fransen, F., et al., The Impact of Gut Microbiota on Gender-Specific Differences in Immunity. Front Immunol, 2017. 8: pp. 754.

52. Biedermann, L., et al., Smoking cessation induces profound changes in the composition of the intestinal microbiota in humans. PLoS One, 2013. 8(3): p. e59260.

53. Pajunen, P., et al., The validity of the Finnish Hospital Discharge Register and Causes of Death Register data on coronary heart disease. Eur J Cardiovasc Prev Rehabil, 2005. 12(2): p. 132–7.

54. Tolonen, H., et al., The validation of the Finnish Hospital Discharge Register and Causes of Death Register data on stroke diagnoses. Eur J Cardiovasc Prev Rehabil, 2007. 14(3): p. 380–5.

55. Sund, R., Quality of the Finnish Hospital Discharge Register: a systematic review. Scand J Public Health, 2012. 40(6): p. 505–15.

56. Hillmann, B., et al., Evaluating the Information Content of Shallow Shotgun Metagenomics. mSystems, 2018. 3(6).

57. Köster, J. and S. Rahmann, Snakemake--a scalable bioinformatics workflow engine. Bioinformatics, 2012. 28(19): p. 2520–2.

58. Kim, D., et al., Centrifuge: rapid and sensitive classification of metagenomic sequences. Genome Res, 2016. 26(12): p. 1721–1729.

