## Supplementary material for "The gut microbiome is a significant risk factor for future chronic lung disease": Fig S1

**Fig S1. A-B**, Prediction of incident asthma and COPD using microbial features at different taxonomic levels individually and in combination. Models were developed in 70% of samples and tested in the rest 30% of samples. **C-D**, Prediction of incident asthma and COPD using gut microbial features at different taxonomy levels using 10 resampled data partitions. Error bars represent mean and standard deviation.

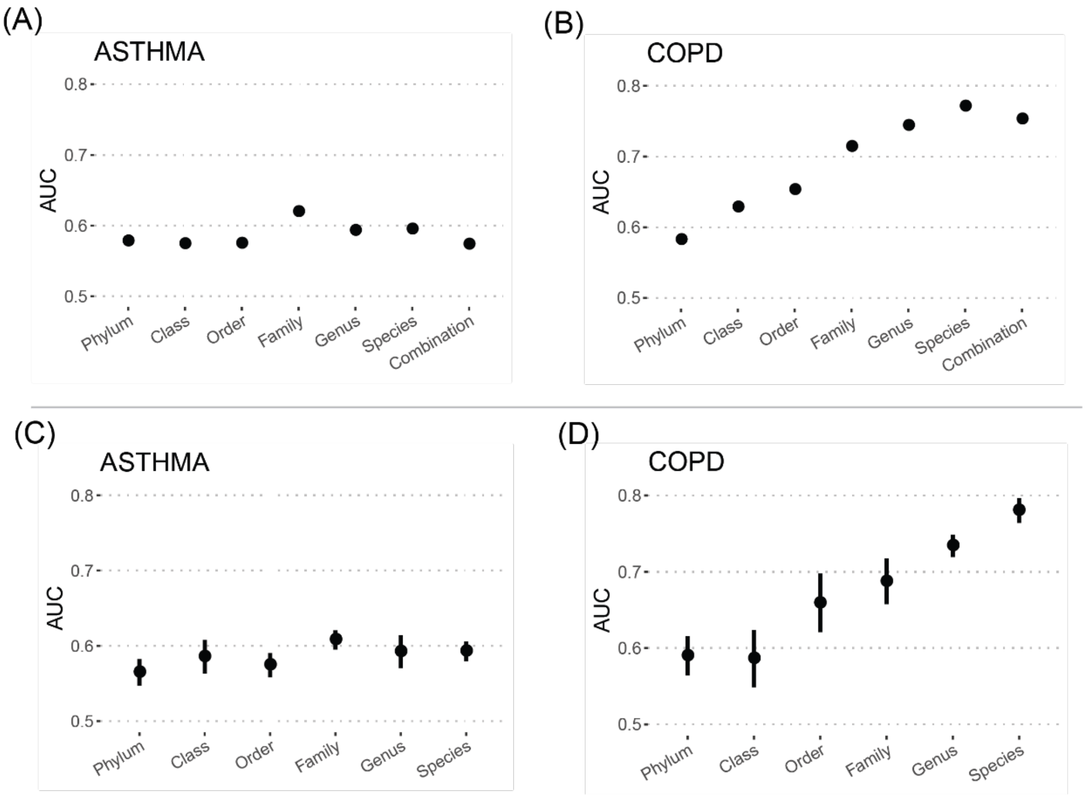
